# Amyloid age and tau PET timeline to symptomatic Alzheimer’s disease in Down syndrome

**DOI:** 10.1101/2024.08.08.24311702

**Authors:** Emily K. Schworer, Matthew D. Zammit, Jiebiao Wang, Benjamin L. Handen, Tobey Betthauser, Charles M. Laymon, Dana L. Tudorascu, Annie D. Cohen, Shahid H. Zaman, Beau M. Ances, Mark Mapstone, Elizabeth Head, William E. Klunk, Bradley T. Christian, Sigan L. Hartley, Alzheimer Biomarker Consortium – Down Syndrome (ABC-DS)

## Abstract

**Background:** Adults with Down syndrome (DS) are at risk for Alzheimer’s disease (AD). Recent natural history cohort studies have characterized AD biomarkers, with a focus on PET amyloid-beta (Aβ) and PET tau. Leveraging these well-characterized biomarkers, the present study examined the timeline to symptomatic AD based on estimated years since reaching Aβ+, referred to as “amyloid age”, and in relation to tau in a large cohort of individuals with DS.

**Methods:** In this multicenter cohort study, 25 – 57-year-old adults with DS (n = 167) were assessed twice from 2017 to 2022, with approximately 32 months between visits as part of the Alzheimer Biomarker Consortium - Down Syndrome. Adults with DS completed amyloid and tau PET scans, and were administered the modified Cued Recall Test and the Down Syndrome Mental Status Examination. Study partners completed the National Task Group-Early Detection Screen for Dementia.

**Findings:** Mixed linear regressions showed significant quadratic associations between amyloid age and cognitive performance and cubic associations between amyloid age and tau, both at baseline and across 32 months. Using broken stick regression models, differences in mCRT scores were detected beginning 2.7 years following Aβ+ in cross-sectional models, with an estimated decline of 1.3 points per year. Increases in tau began, on average, 2.7 – 6.1 years following Aβ+. On average, participants with mild cognitive impairment were 7.4 years post Aβ+ and those with dementia were 12.7 years post Aβ+.

**Interpretation:** There is a short timeline to initial cognitive decline and dementia from Aβ+ (Centiloid = 18) and tau deposition in DS relative to late onset AD. The established timeline based on amyloid age (or equivalent Centiloid values) is important for clinical practice and informing AD clinical trials, and avoids limitations of timelines based on chronological age. **Funding.** National Institute on Aging and the National Institute for Child Health and Human Development.

**Research in Context:** *Evidence before this study:* We searched PubMed for articles published involving the progression of Aβ and tau deposition in adults with Down syndrome from database inception to March 1, 2024. Terms included “amyloid”, “Down syndrome”, “tau”, “Alzheimer’s disease”, “cognitive decline”, and “amyloid chronicity,” with no language restrictions. One previous study outlined the progression of tau in adults with Down syndrome without consideration of cognitive decline or clinical status. Other studies reported cognitive decline associated with Aβ burden and estimated years to AD symptom onset in Down syndrome. Amyloid age estimates have also been created for older neurotypical adults and compared to cognitive performance, but this has not been investigated in Down syndrome.

*Added value of this study:* The timeline to symptomatic Alzheimer’s disease in relation to amyloid, expressed as duration of Aβ+, and tau has yet to be described in adults with Down syndrome. Our longitudinal study is the first to provide a timeline of cognitive decline and transition to mild cognitive impairment and dementia in relation to Aβ+.

*Implications of all the available evidence:* In a cohort study of 167 adults with Down syndrome, cognitive decline began 2.7 – 5.4 years and tau deposition began 2.7 – 6.1 years following Aβ+ (Centiloid = 18). Adults with Down syndrome converted to MCI after ~7 years and dementia after ~12-13 years of Aβ+. This shortened timeline to AD symptomology from Aβ+ and tau deposition in DS based on amyloid age (or corresponding Centiloid values) can inform clinical AD intervention trials and is of use in clinical settings.

---

Individuals with Down syndrome (DS) have a 75-90% lifetime risk of symptomatic Alzheimer’s disease (AD),^1,2^ driven by the overexpression of the amyloid precursor protein (APP) gene on chromosome 21.^3,4^ PET imaging with [C-11]PiB has shown that the accumulation of extracellular brain Aβ plaques in DS can begin in the 30s.^5^ Deposition of Aβ in adults with DS is first most systematically detected in the striatum,^5–7^ similar to autosomal-dominant AD [ADAD] involving APP or presenilin 1 and 2 carriers.^8^ Thereafter, spatial progression of Aβ in DS^5,9^ largely mirrors sporadic late-onset AD [LOAD].^10^ Subsequent to Aβ-positivity [Aβ+], neurofibrillary tau deposition is often observed, following the conventional Braak staging of tau pathology,^11^ similar to LOAD and ADAD.^12–14^

To facilitate therapeutic AD trials in DS and inform clinical decisions, the timeline to symptomatic AD based on Aβ and tau PET neuropathology needs to be established. Initial work evaluated AD biomarkers in relation to estimated years to symptom onset (EYO) by subtracting an individual with DS’s chronological age from the population-mean age of symptomatic AD onset, set at 52.5 years.^1,15^ Timelines based on EYO, however, do not account for important within-population variability in the age of onset of symptomatic AD, which ranges from 45-58 years old.^1^ The range in AD symptom onset is related to heterogeneity in the age of Aβ+ across individuals with DS, shown to span from age 36 to 55 years old.^6,7,16^ This heterogeneity in age of Aβ+ limits the utility of AD timelines based on EYO. To address these limitations, recently, AD timelines centered on Aβ+ chronicity, or “amyloid age” have been created for older neurotypical adults with a family history of AD^17,18^ and individuals with DS.^19^

Amyloid age estimates were created from longitudinal data and based on trajectory modeling that predicts the number of years that an individual has been Aβ+.^17,19^ In our prior DS work, the amyloid age estimate had robust associations with the timing of PET tau deposition,^19^ with increases in tau within the first 2.5 – 5 years of becoming Aβ+. The timing of AD symptomology in relation to amyloid age, and relative to tau burden, remains unknown in DS.

Prior DS studies show associations between PET Aβ and AD-related cognitive impairments.^20,21^ Across three years, increases in PET Aβ predicted declines in memory, executive functioning, and motor processing speed prior to dementia-onset in DS.^22,23^ Adults with DS who were Aβ+ at study onset had greater memory decline relative to those who were Aβ- or who became Aβ+ during the study.^20^ Among individuals with DS who were Aβ+, cognitive declines were only evident with elevated tau, suggesting a short time lag between tau deposition and AD symptomology.^24,25^

The present study sought to establish the timeline to symptomatic AD in relation to amyloid age and relative to tau deposition in DS. Analyses included 167 adults with DS in the Alzheimer Biomarker Consortium - Down Syndrome (ABC-DS) who underwent cognitive testing and neuroimaging at two data collection cycles, spaced 32 months (SD = 6.30) apart. The central hypothesis was that after reaching Aβ+, cognitive decline would be closely tied to the timing of tau deposition, with the transition to MCI and dementia shortly following.

## Methods

### Procedures

Procedures were approved by a central IRB and consent was obtained prior to study activities. Data was collected at four research sites (University of Wisconsin-Madison [UWM], University of Pittsburgh [PITT], University of Cambridge [CAM], and Washington University-St. Louis [WSL]) in the ABC-DS. ^26^ Participants completed brain imaging scans and were administered a cognitive battery at baseline and 32 months later (cycle 2). A study partner completed informant-reports about the participant’s functioning, behavior, and medical history.

### Participants

Analyses included 167 adults with DS (n = 92 with longitudinal data). Inclusion criteria included: 1) ≥ 25 years old, 2) mental age of ≥ 3 years, and 3) trisomy 21 (full, mosaic, or translocation) confirmed through karyotyping. Exclusion criteria included an untreated/unstable medical or psychiatric condition that impaired cognition or a condition contraindicative for an MRI (e.g., metallic implants). Table 1 shows participant demographics.

**Table 1.** Descriptive statistics at baseline.

|  | M (SD) or n (%) |  |  |  |  |
| --- | --- | --- | --- | --- | --- |
|  | Total<br>(n = 167) | CS<br>(n = 141) | MCI<br>(n = 8) | AD<br>(n = 7) | Unable to<br>Determine<br>(n = 11) |
| Age | 38.91 (8.47) | 37.16 (7.41) | 48.88 (5.08) | 52.00 (3.83) | 45.73 (10.13) |
| Aβ+, n (%) | 56 (33.50%) | 36 (25.53%) | 7 (87.50%) | 7 (100%) | 6 (54.55%) |
| Amyloid age (years) | -2.05 (7.06) | -3.70 (5.64) | 7.40 (6.58) | 12.72 (5.61) | 2.89 (6.66) |
| NFTI.II | 1.19 (.21) | 1.14 (0.14) | 1.56 (0.28) | 1.52 (0.30) | 1.35 (0.34) |
| NFT III.IV | 1.14 (.21) | 1.09 (0.09) | 1.51 (0.32) | 1.64 (0.45) | 1.26 (0.27) |
| NFT V.VI | 1.10 (.22) | 1.05 (0.08) | 1.44 (0.40) | 1.66 (0.60) | 1.17 (0.18) |
| Sex, male n (%) | 85 (50.90%) | 69 (48.94%) | 6 (75.00%) | 4 (57.14%) | 6 (54.55%) |
| Premorbid ID, n (%) |  |  |  |  |  |
| Mild | 92 (55.09%) | 78 (55.32%) | 5 (62.50%) | 4 (57.14%) | 5 (45.45%) |
| Moderate | 53 (31.74%) | 42 (29.79%) | 3 (37.50%) | 3 (42.86%) | 5 (45.45%) |
| Severe/Profound | 22 (13.17%) | 21 (14.89%) | - | - | 1 (9.10%) |
| Karyotype, n (%) |  |  |  |  |  |
| Trisomy 21 | 148 (88.62%) | 123 (87.23%) | 7 (87.50%) | 7 (100%) | 11 (100%) |
| Mosaicism | 5 (3.00%) | 4 (2.84%) | 1 (12.50%) | - | - |
| Translocation | 13 (7.78%) | 13 (9.22%) | - | - | - |
| Not available | 1 (.60%) | 1 (.71%) | - | - | - |
| Ethnicity, n (%) |  |  |  |  |  |
| Not Hispanic/Latino | 164 (98.20%) | 138 (97.87%) | 8 (100%) | 7 (100%) | 11 (100%) |
| Hispanic Latino | 3 (1.80%) | 3 (2.13%) | - | - | - |
| mCRT Total score | 31.58 (6.72) | 33.21 (4.13) | 22.25 (9.29) | 18.71 (9.46) | 23.57 (12.71) |
| mCRT Intrusions | 4.56 (5.53) | 3.36 (3.60) | 10.38 (5.85) | 15.71 (9.76) | 10.00 (10.75) |
| DSMSE | 64.30 (12.53) | 66.25 (11.18) | 59.50 (9.79) | 46.50 (12.04) | 49.14 (17.12) |
| NTG-EDSD 6-domain | 3.38 (6.10) | 1.40 (2.33) | 8.38 (5.01) | 19.00 (8.69) | 16.55 (8.41) |

CS = Cognitively stable; MCI = Mild Cognitive Impairment; AD = Alzheimer's disease; Aβ = amyloid-beta; ID = Intellectual disability; mCRT = Modified Cued Recall Test; DSMSE = Down Syndrome Mental Status Examination; NTG-EDSD = National Task Group-Early Detection Screen for Dementia

### Sociodemographic Measures

Study partners reported participant age, biological sex, race, and ethnicity. Apolipoprotein E (APOE) e4 status (present or absent) was determined through genetic testing. Premorbid intellectual disability level (ID) was estimated using the Stanford-Binet, fifth edition abbreviated battery IQ and coded mild, moderate, or severe/profound.

### Cognitive Functioning

*Down Syndrome Mental Status Examination (DSMSE).*^27^ The DSMSE has demonstrated clinical utility for distinguishing adults with DS with versus without AD dementia.^28^ Scores range from 0 to 87 with higher scores indicating better cognitive performance.

*Modified Cued Recall Test (mCRT).*^29^ The mCRT measures episodic memory and is sensitive to AD dementia in DS.^30^ Total scores range from 0 to 36, with higher scores indicating better memory. The mCRT intrusion score specifies the number of incorrect items (i.e., memory errors).

*National Task Group-Early Detection Screen for Dementia (NTG-EDSD).*^31^ The NTG-EDSD is an informant-report that assesses functional and behavioral dementia-related changes. The 6-domain total score (ranges 0 to 51) was used, with higher values indicative of more dementia symptoms. The NTG-EDSD is an accurate screen for MCI (AUC = .76) and dementia (AUC = .94) in DS.^32^

### Clinical Status

Clinical status (cognitively stable, mild cognitive impairment [MCI] or dementia) was determined from a consensus process independent of imaging results.^26^ Cognitive and informant-measure scores were reviewed along with medical and psychiatric histories. If cognitive and/or functional declines were observed but medical or psychiatric conditions or life changes could not be ruled out as the cause, a status of ‘unable to determine’ was given.

### Imaging acquisition and analysis

T1-weighted magnetic resonance imaging (MRI) scans were completed on a GE Signa 750 (UWM), Siemens Trio or Prisma (PITT; WSL), or GE SIGNA PET/MR (CAM) and processed using FreeSurfer v5.3.0. PET scans were performed on a Siemens ECAT HR+ scanner (UWM; PITT), Siemens 4-ring Biograph mCT (UWM; PITT; WSL), and GE Signa PET/MR (CAM). [C-11]PiB (15 mCi) was injected intravenously, and scans were acquired after 50-70 minutes. Standardized uptake value ratio (SUVR) images were generated using gray matter cerebellum as the reference. Following the PiB scan, 10 mCi of [F-18]AV-1451 were injected intravenously, and measurements were acquired after 80-100 minutes.

Amyloid burden was initially quantified by the amyloid load metric and equivalent Centiloids.^33^ Our previous work established a publicly available population-based trajectory of Aβ increase for individuals with DS using a sampled iterative local approximation (SILA) algorithm.^19^ Briefly, longitudinal Aβ PET trajectories with respect to chronological age were modeled using the Euler method to generate a population-averaged curve of Aβ Centiloids with respect to time, denoted as Aβ chronicity or “amyloid age.” The amyloid age curve is designed such that an age of zero years represents the onset of PET amyloid-positivity, which has been defined as 18.0 Centiloids for this population.^16^ Then, for each individual with DS, their Aβ Centiloid values were aligned to this curve to determine the amyloid age at that scan. The amyloid age was then subtracted from the participants chronological age to determine the estimated years to/from Aβ+. A sampled iterative local approximation (SILA) algorithm was used to model Aβ trajectories longitudinally from the PiB scan and assign each participant an amyloid age value, representing the duration of Aβ+ in years. Amyloid age values were centered at 18 equivalent Centiloids (Aβ+ = 0 years).^19^

### Analysis plan

The distributions for amyloid age, tau PET SUVR, and cognitive variables were examined for skewness and outliers. Mixed generalized linear models examined the association between amyloid age (considering up to cubic polynomials) and mCRT, DSMSE, and NTG-EDSD at baseline, adjusting for sex, premorbid ID, and APOE e4 status. For participants with longitudinal data, cognitive change scores were created (cycle 2 minus baseline scores). Mixed generalized linear models evaluated the association between amyloid age and cognitive change scores with the same covariates described above. Next, mixed generalized linear models examined the association between amyloid age and tau PET SUVR in Braak NFT regions I-II, III-IV, V-VI, controlling for sex, premorbid ID, and APOE e4 status for baseline and longitudinal data. In models, the mcp R package^34^ identified the amyloid age value that corresponded to decreases in cognitive performance and increases in tau PET following Aβ+ with broken stick regression. This approach assumes the slope before the change point is zero (plateau), and uses a Bayesian approach to identify change points and model the regression function flexibly. A nonzero after-changepoint slope is considered if its 95% credible interval does not cover zero. Finally, amyloid age and tau PET were compared across AD clinical statuses using one-way between subjects ANOVA with Tukey’s HSD.

## Results

There were no significant differences in the distribution of sex, race, ethnicity, APOE e4 status, DS type, or premorbid ID for the participants with baseline-only data compared to those with longitudinal data (*p* > 0.05). During the study, three (3.3%) participants converted from cognitively stable to MCI, one (1.1%) converted from cognitively stable to dementia, and five (5.4%) converted from MCI to dementia. Amyloid age and DSMSE scores were normally distributed. There was slight skewness for the mCRT total (skew = −2.3), mCRT intrusions (skew = 2.1), NTG-EDSD (skew = 2.8), and tau PET SUVR (skew = 1.7 – 4.2), which was expected given that most participants were cognitively stable. Of the 167 participants, 56 (33.5%) were Aβ+ at baseline.

Mixed model linear regressions examined the effect of baseline amyloid age on baseline cognitive performance controlling for covariates. There was a significant association between premorbid ID and mCRT and DSMSE scores. Participants with mild ID had higher mCRT and DSMSE scores than those with moderate or severe ID. There was also a significant association between sex and baseline DSMSE scores; females scored 3.3 points higher than males, on average. APOE e4 status was not a significant predictor in any models. There was a quadradic association between amyloid age and mCRT total, R^2^ = .48, *F*(6, 150) = 23.23, *p* < .01, mCRT intrusions, R^2^ = .44, *F*(6, 150) = 19.32, *p* < .01, DSMSE, R^2^ = .52, *F*(6, 152) = 27.67, *p* < .01, and NTG-EDSD, R^2^ = .25, *F*(6, 158) = 8.76, *p* < .01. In models of longitudinal cognitive change, there was also a significant quadratic effect of amyloid age on cognitive change, for all outcomes except mCRT intrusions (Figure 1; Supplementary Table 1). Figure 1 shows Centiloid values for direct comparison with amyloid age.

**Figure 1.**
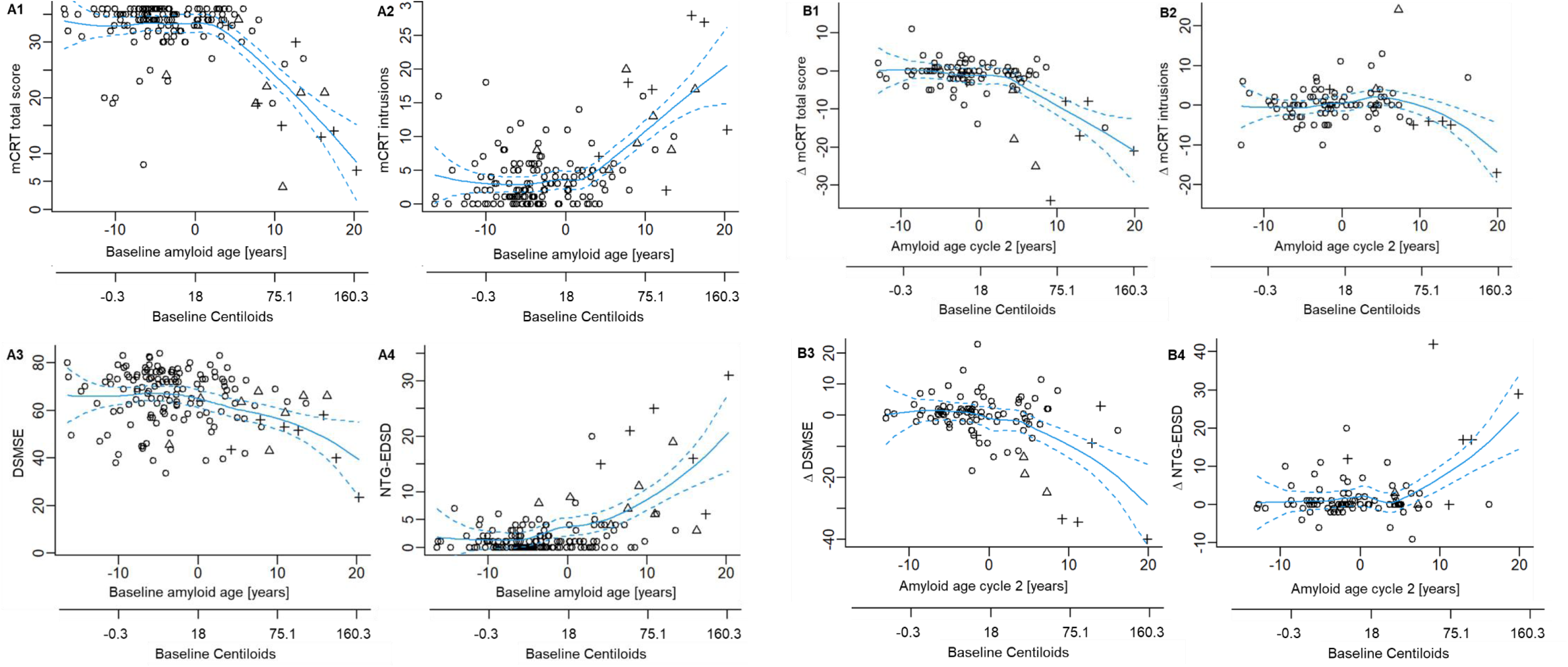
**A)** Scatterplots and Loess visualizations of the association between amyloid age and cognitive performance at baseline; n = 167; ○ = cognitively stable, Δ = MCI, + = AD. Amyloid age of 0 years indicates Aβ+ (18 CL). **B)** Scatterplots and Loess visualizations of the association between amyloid age and change in cognitive domains (across ~32 months); n = 92; Δ mCRT, DSMSE, and NTG-EDSD indicates change from baseline to cycle 2.

Mixed model linear regressions examined the effect of baseline amyloid age on baseline tau PET SUVR in NFT regions I-II, III-IV, V-VI controlling for covariates. There was a quadradic association between amyloid age and NFT regions I-II, R^2^ = .64, *F*(6, 159) = 46.78, *p* < 0.01. There was a cubic association between amyloid age and NFT regions III-IV, R^2^ = .77, *F*(7, 158) = 76.27, *p* < 0.01 and NFT regions V-VI, R^2^ = .75, *F*(7, 158) = 66.99, *p* < 0.01. Models predicting tau change were cubic associations between amyloid age and change in NFT regions I-II, R^2^ = .29, *F*(7, 84) = 4.78, *p* < 0.01, NFT regions III-IV, R^2^ = .32, *F*(7, 84) = 5.51, *p* < .01 and NFT regions V-VI, R^2^ = .28, *F*(7, 84) = 4.64, *p* < .01 (Figure 2; Supplementary Table 2). Figure 2 includes Centiloid values for direct comparison with amyloid age.

**Figure 2.**
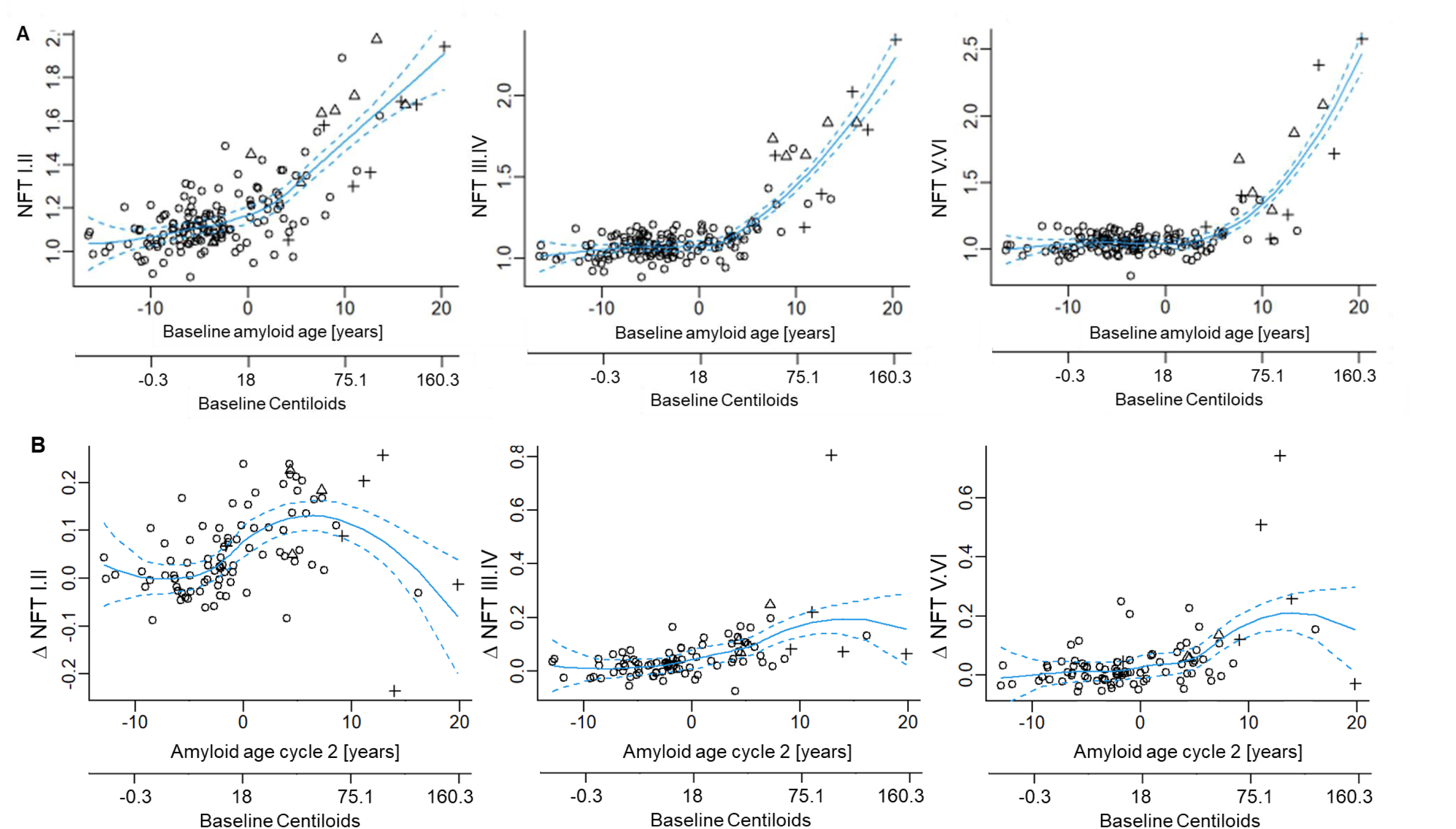
**A)** Scatterplots and Loess visualizations of the association between amyloid age and tau (NFT I.II; NFT III.IV; NFT V.VI) at baseline; ○ = cognitively stable, Δ = MCI, + = AD. Amyloid age of 0 years indicates Aβ+ (18 CL). **B)** Scatterplots and Loess visualizations of the association between amyloid age and change in tau (NFT I.II; NFT III.IV; NFT V.VI) at baseline. Δ NFT indicates change from baseline to cycle 2.

Broken stick regressions identified change points after Aβ+ for cognitive measures (Table 2). In models predicting baseline performance, following Aβ+, there was an average change point of 2.7 years for the mCRT total, 2.8 years for mCRT intrusions, and 3.8 years for NTG-EDSD. After these change points, mCRT total decreased by 1.3 points, mCRT intrusions increased by 1.0 point, and NTG-EDSD increased by 0.8 points per year. Change points after Aβ+ were also identified for tau deposition relative to amyloid age. There was an average change point of 2.7 years for NFT regions I-II, 3.4 years for NFT regions III-IV, and 6.1 years for NFT regions V-VI. After these change points, tau PET SUVR increased by 0.04 – 0.1 units per year.

**Table 2.** Broken stick regression identifying change point in cognitive performance and tau in relation to amyloid age for individuals with amyloid age values ≥ 0.

| Cognitive functioning vs. amyloid age (baseline) |  |  |  | Slope after the change point |  |  |
| --- | --- | --- | --- | --- | --- | --- |
|  | **Change point** | **Lower** | **Upper** | **Mean** | **Lower** | **Upper** |
| mCRT total score | **2.68** | 0.21 | 5.40 | -1.33 | -1.83 | -0.88 |
| mCRT intrusions | **2.76** | 0.21 | 5.40 | 0.99 | 0.60 | 1.40 |
| DSMSE | 8.51 | 0.22 | 17.70 | -1.12 | -3.55 | 0.88 |
| NTG-EDSD 6-domain | **3.80** | 0.21 | 8.31 | 0.80 | 0.28 | 1.34 |
| NFT I.II | **2.71** | 0.24 | 4.99 | 0.04 | 0.03 | 0.05 |
| NFT III.IV | **3.38** | 1.44 | 5.14 | 0.06 | 0.05 | 0.07 |
| NFT V.VI | **6.11** | 3.18 | 10.50 | 0.08 | 0.05 | 0.14 |
| Cognitive change score vs. amyloid age (cycle 2) |  |  |  | Slope after the change point |  |  |
|  | **Change point** | **Lower** | **Upper** | **Mean** | **Lower** | **Upper** |
| $\Delta$ mCRT total score | **3.56** | 0.03 | 8.97 | -0.98 | -1.73 | -0.24 |
| $\Delta$ mCRT intrusions | 8.63 | 0.05 | 18.00 | -0.46 | -1.34 | 0.39 |
| $\Delta$ DSMSE | 6.54 | 0.04 | 16.40 | -1.05 | -2.50 | 0.45 |
| $\Delta$ NTG-EDSD 6-domain | **5.43** | 0.03 | 12.80 | 1.03 | 0.01 | 2.08 |
| $\Delta$ NFT I.II | 9.04 | 0.52 | 18.80 | -0.01 | -0.02 | 0.01 |
| $\Delta$ NFT III.IV | 7.57 | 0.03 | 18.30 | 0.01 | -0.01 | 0.02 |
| $\Delta$ NFT V.VI | 5.69 | 0.03 | 17.20 | 0.01 | -0.01 | 0.02 |

*Note.* Significant nonzero slope based on 95% credible interval after the change point bolded. mCRT = Modified Cued Recall Test; DSMSE = Down Syndrome Mental Status Examination; NTG-EDSD = National Task Group-Early Detection Screen for Dementia; $\Delta$ indicates change from baseline to cycle 2.

In models predicting change in cognitive measures from baseline to cycle 2, there was a change point of 3.6 years for the mCRT total. After this change point, the mCRT total decreased by 1.0 point per year. There was a significant change point for the NTG-EDSD at 5.4 years, with scores increasing by 1.0 per year thereafter (Table 2). No significant change slopes were identified post Aβ+ in NFT regions I-VI from baseline to cycle 2.

At baseline there was a significant effect of AD clinical status on amyloid age, *F*(3, 163) = 29.19, *p* < 0.01. Cognitively stable participants had an average amyloid age of −3.7, *SD* = 5.6, and had a lower amyloid age value than participants with MCI (*M* = 7.4, *SD* = 6.6) or dementia (M = 12.7, SD = 5.6), *p* < 0.01. There was no significant difference in amyloid age between participants with MCI versus dementia (*p* = .28; Supplementary Figure 1). Participants with an ‘unable to determine’ status had significantly higher amyloid age (*M* = 2.9, *SD* = 6.7) than cognitively stable participants but lower than participants with dementia (*p* < 0.01).

At baseline there was a significant effect of AD clinical status on tau PET SUVR NFT regions I-II, *F*(3, 163) = 26.70, *p* < 0.01, NFT regions III-IV, *F*(3, 163) = 47.13, *p* < 0.01, and NFT regions V-VI, *F*(3, 163) = 42.32, *p* < 0.01 (Supplementary Figure 2). Across all regions, cognitively stable participants had lower tau PET than those with MCI and dementia (*p* < 0.01). There were no significant differences between participants with MCI and dementia (*p* = 0.06 – 0.96). Participants with an ‘unable to determine’ status had significantly higher tau PET in NFT regions I-IV than cognitively stable participants (*p* < 0.01). Tau PET in NFT regions III-VI was significantly lower in the ‘unable to determine’ group compared to MCI and dementia groups (*p* < 0.01). Figure 3 shows the timeline of tau, cognitive decline, and MCI and dementia based on amyloid age.

**Figure 3.**
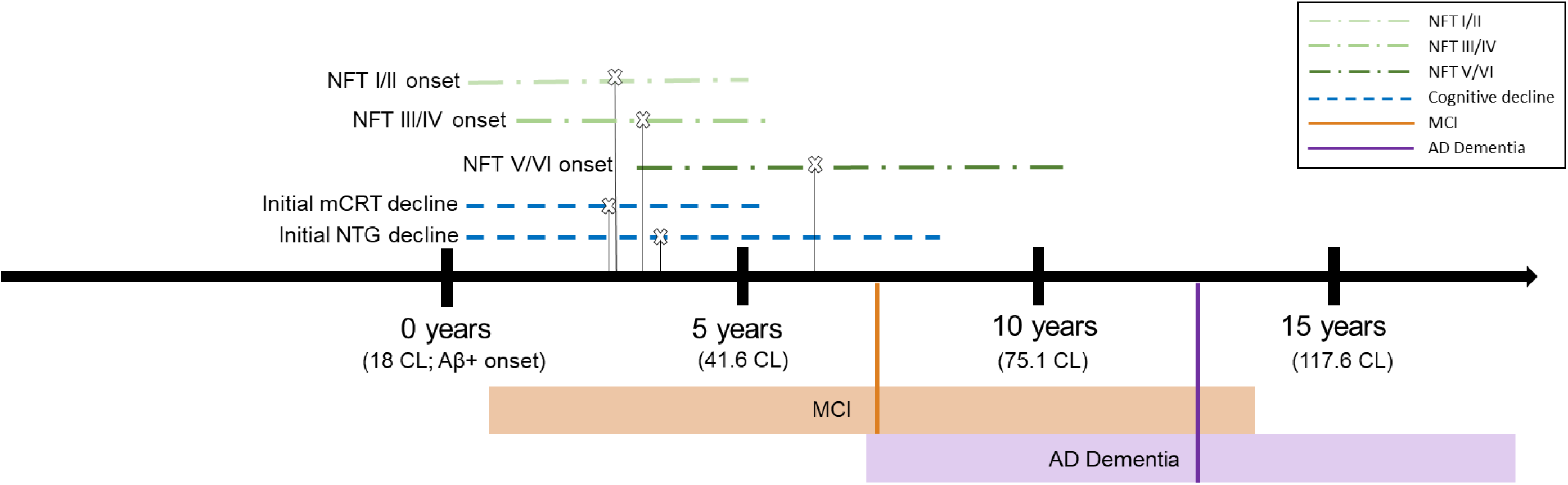
Timeline to symptomatic Alzheimer’s Disease in Down syndrome. Mean change points for cognitive decline (mCRT and NTG-EDSD) and tau (Braak NFT regions I-VI) marked with **x**. Cognitive decline and tau upper and lower ranges indicated with blue and green dotted lines. Average amyloid age values for MCI and AD dementia marked with orange and purple lines respectively. Shaded orange and purple bars show standard deviations. CL = Centiloids; mCRT = Modified Cued Recall Test; NTG = National Task Group-Early Detection Screen for Dementia

## Discussion

The present study describes the timeline of symptomatic AD relative to duration of Aβ+ (“amyloid age”) in DS. Amyloid age can be directly related to Centiloid magnitude, and thus this timeline is of high clinical utility for AD intervention trials and practice. Findings indicate that, cognitive performance initially remains stable for the first 2-3 years after becoming Aβ+ before declining. Following this stable period (2.7 years based on baseline and 3.6 years based on change scores), mCRT scores decreased by 1.3 and 1.0 points per year, respectively. For mCRT intrusions, a change point was identified at 2.8 years post Aβ+, with an increase of 1.0 intrusions per year thereafter. Changes in the NTG-ESDS did not begin until 3.8 (baseline) or 5.4 (cycle 2) years following Aβ+. The one-to-two-year lag in change on the NTG-ESDS, relative to mCRT, mirrors prior reports that episodic memory is the first cognitive domain affected in AD in DS^23,35,36^ and that direct measures are more sensitive to early AD-related declines than informant-reports.^32^

The change points from the within-person longitudinal analyses, which are generally viewed as more robust than cross-sectional estimates, can guide longitudinal clinical AD intervention design. Efficacious interventions involving the mCRT total as an outcome would be expected to demonstrate delayed onset of decline after Aβ+ (>3.6 years) and/or a slowed rate of decline (<1.3 points/year) relative to this natural history cohort. This study also offers meaningful information for the timeline for entering individuals with DS into clinical AD interventions. Intervention effects may be optimized for individuals with DS who have not yet reached Aβ+ or within the first 3 years of being Aβ+, as cognitive decline has not yet begun.

There was a cubic pattern of association between tau deposition and amyloid age. Change points in tau deposition were identified only in models using baseline data. Following Aβ+, initial increases in tau deposition occurred at 2.7[0.2,5.0] years in NFT I-II, 3.4[1.4,5.1] years in NFT III-IV, and 6.1[3.2,10.5] years in NFT V-VI. The timing of tau deposition in NFT I-II is closely aligned with initial cognitive decline in DS. Declines in NFT V-VI lag an estimated 3 years from initial medial temporal tau deposition, consistent with our previous findings.^19^ When evaluating change in NFT, an initial rapid increase in tau burden followed Aβ+ before reaching a plateau, or, in the case of NFT I-II, decrease at high amyloid age (≥ 15 years or Centiloid = 117.6). In DS, ventricle enlargement is common with aging and AD, which erodes the NFT I-II ROIs and introduces partial volume effects. The change observed in NFT regions III-IV and V-VI show greater uncertainty at high amyloid ages due to the small sample size in this range. More accurate trajectories at these high amyloid ages (or Centiloid values) should be modeled as ABC-DS progresses.

Similar to previous findings,^19,22,23^ we did not observe effects of sex or APOE e4 on imaging or mCRT and NTG-EDSD outcomes. This distinguishes DS from LOAD, where individuals with APOE e4 allele and women exhibit higher risk of AD pathology.^37^ Other DS studies have identified effects of APOE e4 on AD biomarker onset,^38,39^ in which dementia occurs ~2 years earlier.^40^ Given the young age of our cohort, and low incidence of MCI and dementia, we may be evaluating biomarker change too early to capture APOE effects.

The current study is the first to report the timing of MCI and dementia relative to amyloid age and in relation to tau burden in DS. Average amyloid age for individuals with DS with MCI was 7.4 years (SD = 6.6) and 12.7 (SD = 5.6) years for those with dementia, corresponding to Centiloid = 62.1 and 99.3 respectively. This suggests an accelerated timeline to AD symptomology in DS relative to LOAD, where progression to MCI occurs 15.5 years post Aβ+.^18^ Individuals with DS with a clinical status of ‘unable to determine’ had a mean amyloid age of 2.9 years. Many of these individuals were likely exhibiting initial AD-related symptoms, matching their biomarker profile -- comparable amyloid age to individuals with MCI and higher tau burden in NFT I-IV than those who were cognitively stable.

Limitations to the current study include a low proportion of individuals with MCI or dementia and that average amyloid age (as opposed to amyloid age at initial transition to these clinical statuses) was evaluated. Thus, the amyloid age values associated with MCI or dementia may be overestimated. In addition, amyloid age estimates that are negative display poor predictive power in determining the onset of Aβ+, directly due to the native signal detection limits of PET scanners. This limitation was mitigated by focusing the regression models to identify inflection points following the onset of amyloid (amyloid age = 0 years, Centiloid = 18.0). Longitudinal models were based on two data collection cycles spanning 3 years; however, longer time frames should be evaluated in future studies. Most participants were White and non-Hispanic, and efforts are needed to increase participation from underrepresented groups. Finally, while DSMSE declines were associated with amyloid age, significant inflection points of change were not detected potentially due to higher between and within-person variability in DSMSE scores. The DSMSE also assesses a wide range of cognitive skills, and decline may occur at a more advanced stage in AD progression.

## Conclusion

This study documents the timeline to AD symptomology in relation to amyloid age and tau in DS. Findings indicate a short time from Aβ+ to initial cognitive decline (3 years) in DS, with declines closely aligned with tau in NFT regions I-II, relative to LOAD.^41,42^ On average, individuals with DS transition to MCI after ~7 years of Aβ+ and dementia after ~12 to 13 years. Our AD symptom timeline based on amyloid age can be directly related to Centiloid magnitude^19^ and thus has utility for AD clinical trials and practice. For example, an adult with DS with PET Centiloid of 31, which equates to an amyloid age of 3, would have an estimated 4 years to MCI and 9 years to AD dementia. Timelines based on amyloid age offer improvements over timelines based on EYO, which do not account for marked within-population variability in age of Aβ+ in DS. The amyloid age estimates used in this study are publicly available^19^ and provide the timeline to AD symptomology without intervention, information needed to design clinical AD intervention trials in DS.

## Funding

The Alzheimer’s Biomarkers Consortium–Down Syndrome (ABC-DS) is funded by the National Institute on Aging and the National Institute for Child Health and Human Development (U01 AG051406, U01 AG051412, U19 AG068054). The work contained in this publication was also supported through the following National Institutes of Health Programs: The Alzheimer’s Disease Research Centers Program (P50 AG008702, P30 AG062421, P50 AG16537, P50 AG005133, P50 AG005681, P30 AG062715, and P30 AG066519), the Eunice Kennedy Shriver Intellectual and Developmental Disabilities Research Centers Program (U54 HD090256, U54 HD087011, and P50 HD105353), the National Center for Advancing Translational Sciences (UL1 TR001873, UL1 TR002373, UL1 TR001414, UL1 TR001857, UL1 TR002345), the National Centralized Repository for Alzheimer Disease and Related Dementias (U24 AG21886), and DS-Connect® (The Down Syndrome Registry) supported by the Eunice Kennedy Shriver National Institute of Child Health and Human Development (NICHD). Research reported in this publication was also supported by NICHD under award number T32 HD007489. In Cambridge, UK this research was supported by the NIHR Cambridge Biomedical Research Centre and the Windsor Research Unit, CPFT, Fulbourn Hospital Cambridge, UK. Sampled iterative local approximation algorithm (SILA) methodology was also supported by the National Institute on Aging (R01AG080766).

## Data Availability

All data used in this study are available from the Alzheimer Biomarker Consortium- Down Syndrome data repository located in the University kf Southern California Image and Data Archive: https://ida.loni.usc.edu/login.jsp

## Acknowledgements

The authors are grateful to the ABC-DS study participants, their families and care providers, and the ABC-DS research and support staff for their contributions to this study. This manuscript has been reviewed by ABC-DS investigators for scientific content and consistency of data interpretation with previous ABC-DS study publications. The content is solely the responsibility of the authors and does not necessarily represent the official views of the NIH, the CPFT, the NIHR or the UK Department of Health and Social Care. AVID Radiopharmaceuticals provided the precursor and reference standard to produce AV-1451.

## Notes

### Competing Interest Statement

The authors have declared no competing interest.

### Funding Statement

This study was funded by the National Institutes of Health

### Author Declarations

The Ethics committee/Institutional Review Board of the University of Wisconsin-Madison and ADVARRA gave ethical approval of this work.

