## Supplementary material for "Amyloid age and tau PET timeline to symptomatic Alzheimer’s disease in Down syndrome": No supplemental file

Supplementary Table 1.

*Regression models predicting cognitive skills and dementia symptoms at baseline/change in cognition and dementia symptoms with amyloid age.*

| | mCRT total score | | | mCRT intrusions | | | DSMSE | | | NTG-EDSD | | |
|---|---|---|---|---|---|---|---|---|---|---|---|---|
| | B | 95% CI | p | B | 95% CI | p | B | 95% CI | p | B | 95% CI | p |
| Amyloid age | -0.49 | -0.60, -0.38 | < 0.01 | 0.41 | 0.32, 0.51 | < 0.01 | -0.61 | -0.81, -0.41 | <0.01 | 0.38 | 0.26, 0.50 | < 0.01 |
| Amyloid age$^2$ | -0.04 | -0.05, -0.03 | < 0.01 | 0.03 | 0.02, 0.04 | < 0.01 | -0.03 | -0.05, -0.01 | <0.01 | 0.02 | 0.01, 0.03 | < 0.01 |
| Sex (female) | 1.01 | -0.61, 2.63 | 0.22 | -1.11 | -2.49, 0.28 | 0.12 | 3.02 | 0.16, 5.89 | 0.04 | 0.20 | -1.51, 1.91 | 0.82 |
| Lifetime ID level (vs. Mild) | | | | | | | | | | | | |
| Moderate | -1.32 | -3.11, 0.48 | 0.15 | -0.04 | -1.57, 1.50 | 0.96 | -8.99 | -12.19, -5.80 | < 0.01 | 1.21 | -0.69, 3.12 | 0.22 |
| Severe/Profound | -6.27 | -8.74, -3.79 | < 0.01 | 3.62 | 1.50, 5.75 | < 0.01 | -23.76 | -28.08, -19.45 | < 0.01 | 1.22 | -1.36, 3.80 | 0.35 |
| APOE e4 | 0.99 | -0.95, 2.94 | 0.32 | -0.10 | -1.77, 1.58 | 0.91 | 0.11 | -3.33, 3.56 | 0.95 | -0.60 | -2.67, 1.47 | 0.57 |
| | Δ mCRT total score | | | Δ mCRT intrusions | | | Δ DSMSE | | | Δ NTG-EDSD | | |
| Amyloid age | -0.41 | -0.60, -0.22 | < 0.01 | 0.29 | 0.02, 0.55 | 0.03 | -0.47 | -0.77, -0.18 | < 0.01 | 0.25 | 0.02, 0.48 | 0.03 |
| Amyloid age$^2$ | -0.03 | -0.05, -0.01 | < 0.01 | -0.01 | -0.03, 0.02 | 0.51 | -0.04 | -0.07, -0.01 | 0.01 | 0.04 | 0.01, 0.06 | < 0.01 |
| Amyloid age$^3$ | | | | -0.002 | -0.01, $-3.25 \times 10^{-4}$ | 0.02 | | | | | | |
| Sex (female) | | | | -0.42 | -2.52, 1.69 | 0.70 | | | | | | |
| Lifetime ID level (vs. Mild) | 1.06 | -1.23, 3.34 | 0.36 | | | | 1.22 | -2.38, 4.82 | 0.50 | -0.22 | -2.97, 2.53 | 0.88 |
| Moderate | -0.01 | -2.67, 2.66 | 0.99 | 0.17 | -2.27, 2.61 | 0.89 | -0.61 | -4.80, 3.58 | 0.77 | 0.93 | -2.25, 4.12 | 0.56 |
| Severe/Profound | 0.30 | -2.70, 3.30 | 0.84 | -0.18 | -2.92, 2.57 | 0.90 | 1.19 | -3.53, 5.92 | 0.62 | -0.14 | -3.83, 3.55 | 0.94 |
| APOE e4 | 1.21 | -1.45, 3.87 | 0.37 | -0.64 | -3.07, 1.78 | 0.60 | 2.40 | -1.78, 6.58 | 0.26 | -0.91 | -4.10, 2.28 | 0.57 |

mCRT = Modified Cued Recall Test; DSMSE = Down Syndrome Mental Status Examination; NTG-EDSD = National Task Group-Early Detection Screen for Dementia; ID = Intellectual disability; APOE = Apolipoprotein E

Supplementary Table 2.
*Regression models predicting tau PET SUVR in Braak NFT regions I-II, III-IV, V-VI with amyloid age. ∆ = change in Braak NFT*

| | *Braak NFT regions I-II* | | | *Braak NFT regions III-IV* | | | *Braak NFT regions V-VI* | | |
|---|---|---|---|---|---|---|---|---|---|
| | B | 95% CI | p | B | 95% CI | p | B | 95% CI | p |
| Amyloid age | 0.02 | 0.01, 0.02 | < 0.01 | 0.01 | 0.01, 0.02 | <0.01 | <0.01 | - <0.01, 0.01 | 0.09 |
| Amyloid age$^2$ | 0.01 | <0.01, 0.01 | <0.01 | <0.01 | <0.01, <0.01 | <0.01 | <0.01 | <0.01, <0.01 | <0.01 |
| Amyloid age$^3$ | | | | <0.01 | <0.01, <0.01 | <0.01 | <0.01 | <0.01, <0.01 | <0.01 |
| Sex (female) | -0.01 | -0.04, 0.04 | 0.97 | -0.01 | -0.04, 0.03 | 0.78 | - 0.01 | -0.04, 0.03 | 0.78 |
| Lifetime ID level (vs. Mild) | | | | | | | | | |
| Moderate | 0.03 | -0.02, 0.07 | 0.21 | <0.01 | -0.03, 0.04 | 0.87 | -0.02 | -0.06, 0.02 | 0.29 |
| Severe/Profound | -0.01 | -0.07, 0.05 | 0.69 | -0.01 | -0.06, 0.04 | 0.58 | - <0.01 | -0.06, 0.05 | 0.94 |
| APOE e4 | 0.02 | -0.03, 0.07 | 0.48 | -0.01 | -0.05, 0.03 | 0.79 | - <0.01 | -0.05, 0.04 | 0.90 |
| | *∆ Braak NFT regions I-II* | | | *∆ Braak NFT regions III-IV* | | | *∆ Braak NFT regions V-VI* | | |
| Amyloid age | 0.01 | 0.01, 0.02 | <0.01 | 0.01 | 0.01, 0.02 | <0.01 | 0.01 | 0.01, 0.02 | <0.01 |
| Amyloid age$^2$ | <0.01 | -<0.01, <0.01 | 0.86 | <0.01 | <0.01, <0.01 | 0.01 | <0.01 | <0.01, <0.01 | 0.01 |
| Amyloid age$^3$ | <0.01 | -<0.01, -<0.01 | <0.01 | -<0.01 | -<0.01, -<0.01 | 0.01 | -<0.01 | -<0.01, -<0.01 | 0.03 |
| Sex (female) | -0.01 | -0.04, 0.03 | 0.68 | -<0.01 | -0.04, 0.03 | 0.87 | -<0.01 | -0.05, 0.04 | 0.88 |
| Lifetime ID level (vs. Mild) | | | | | | | | | |
| Moderate | <0.01 | -0.04, 0.04 | 0.85 | 0.03 | -0.02, 0.07 | 0.27 | 0.02 | -0.03, 0.07 | 0.50 |
| Severe/Profound | 0.01 | -0.04, 0.05 | 0.67 | <0.01 | -0.05, 0.05 | 0.95 | -0.01 | -0.06, 0.05 | 0.81 |
| APOE e4 | 0.01 | -0.03, 0.05 | 0.75 | -0.01 | -0.05, 0.04 | 0.78 | -0.01 | -0.06, 0.04 | 0.68 |

ID = Intellectual disability; APOE = Apolipoprotein E

Supplementary Figure 1. ANOVA comparison between amyloid age categorized by clinical status (cognitively stable, MCI, AD, unable to determine). Amyloid age of 0 years indicates Aβ+ (18 CL).

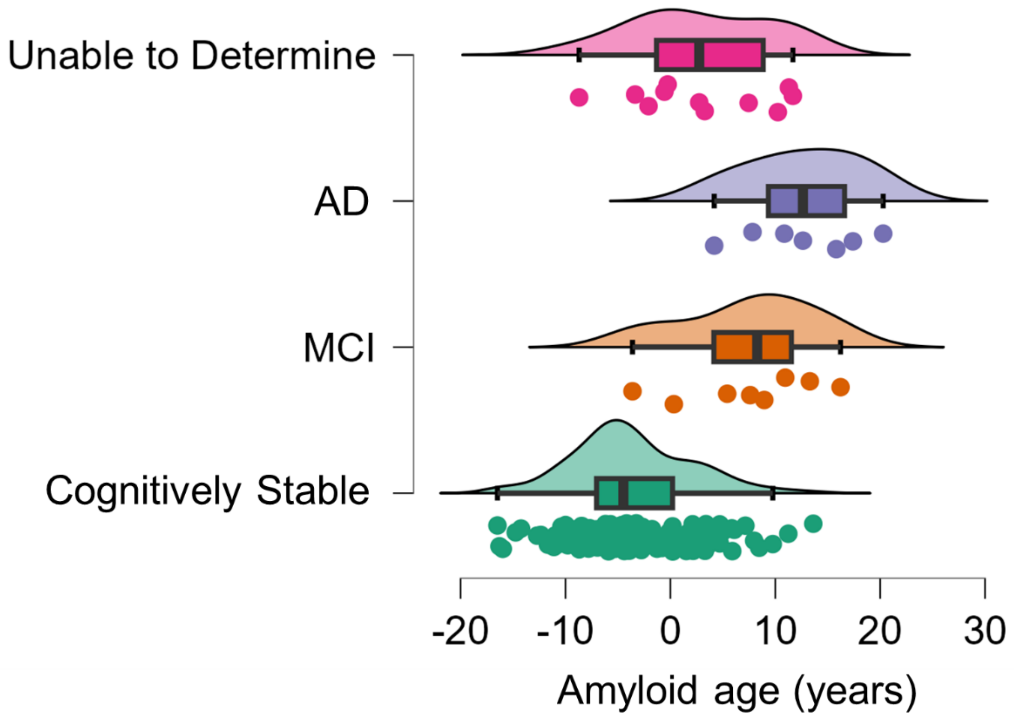

Supplementary Figure 2. ANOVA comparisons between tau Braak NFT SUVR (NFT I.II; NFT III.IV; NFT V.VI) categorized by clinical status (cognitively stable, MCI, AD, unable to determine).

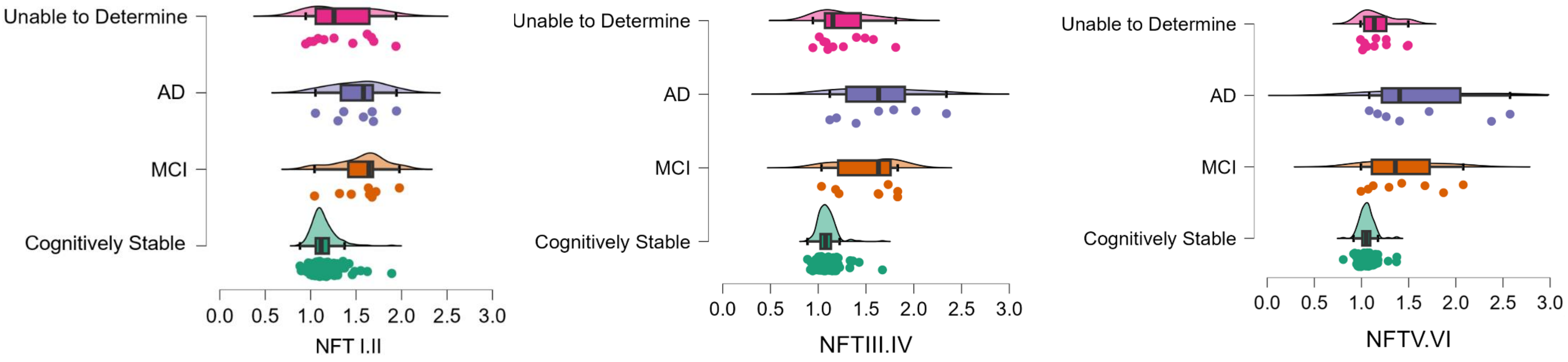
